# Comparative Analysis of Bacteriological and Physico-Chemical Characteristics of Water Samples from Different Sources used by Students of Federal University Dutsin-Ma (FUDMA), Katsina State

**DOI:** 10.1101/2022.04.29.22273784

**Authors:** Ignatius Mzungu, Anastesia Chisom Ebunam

## Abstract

The aim of this study is to comparatively analyze the bacterial load and physiochemical parameters of water samples from various sources used by the students of Federal University, Dutsinma Katsina State. Samples from tap, well, dam, rain, sachet and boreholes were collected at different locations where students reside. There were 6 sources of water, namely; tap water, dam water, well water, borehole water, sachet water and rain water from which 10 samples were obtained each, making a total of 60 samples for analysis. The physicochemical parameters of each water samples were detected. According the technique adopted by Chessbrough. (2000), the samples were serially diluted, 3 test tubes were sterilized and distilled water of 9ml were Pipette into these test tubes, 1ml of the water sample was pipette into the first test tube and was shaken vigorously to have a homogeneous mixture (stock). Bacterial count of each water sample was carried out and presence of *Escherichia coli, P.aeruginosa, S.aureus, S*.typhi, *K.pneumoniae, B.subtilis, Proteus sp, Shigella sp*, and *E.aerogenes* were identified. Biochemical tests were carried out for accurate characterization of the isolates. The pattern of occurrence the studied physico–chemical parameters (except pH) of borehole water, sachet water, Dam, Rain, well, tap water were within the permissible limit set by World Health Organization. The pH of all samples of sachet water were within the permissible limit set by World Health Organization However, the pH of 7 out of 10 samples of borehole water and 8 out of 10 samples of tap water were within the permissible limit set by World Health Organization. The prevalence of indicator organisms in water samples are as follows; *Klebsiella pneumoniae* (Dam water=100%, Sachet water=0, Tap water=80%, Borehole=70%, Rain=20%, Well=100%), *Escherichia coli* (Dam water=100%, Sachet water=0, Tap water=20%, Borehole=10%, Rain=0, Well=100%), *Pseudomonas aeruginosa* (Dam water=100%, Sachet water=60%, Tap water=100%, Borehole=100%, Rain=100, Well=100%), *Staphylococcus aureus* (Dam water=100%, Sachet water=40%, Tap water=50%, Borehole=50%, Rain=20%, Well=100%), *Salmonella* TYPHI (Dam water=100%, Sachet water=0, Tap water=30%, Borehole=10%, Rain=10%, Well=100%), *Bacillus subtilis* (Dam water=100%, Sachet water=60%, Tap water=70%, Borehole=50%, Rain=30%, Well=100%), *Proteus sp* (Dam water=100%, Sachet water=0, Tap water=50%, Borehole=30%, Rain=10%, Well=100%), *Shigella sp* (Dam water=100%, Sachet water=0, Tap water=30%, Borehole=10%, Rain=10%, Well=100%), *Enterobacter aerogenes* (Dam water=100%, Sachet water=30%, Tap water=60%, Borehole=50%, Rain=30%, Well=100%). The research indicates the polluted condition of water in Dutsin-ma. Only sachet water is fit for consumption without further treatment in Dutsin-ma. Tap and borehole water should be treated before consumption. Dam water and well water should be used for other domestic purposes. However it should be treated by sedimentation followed by boiling. Rain water has less bacterial load but has an acidic pH, therefore it is unfit for consumption.

## INTRODUCTION

Water is one of the most important as well as one of the most bountiful of all compounds that is very vital to living organisms (Tortora *et al*., 2002). Water is a chemical compound, it comprises of dual atoms of hydrogen combined with one atom of oxygen. Water is an unscented, tasteless and colorless mobile liquid with the exception of large volumes where it appears blue. It has a melting point of 0°C and a boiling point of 100□ (212□) (Willey *et al*., 2008). It has numerous uses such as washing, food preparation, and food processing, and swimming among others. Drinking of water is the most sensitive out of these uses as it could have a direct harmful effect on health of human beings. Therefore, drinking water should be potable, free from diseases or toxic substances (Pruss *et al*., 2000). Diseases caused by contaminated water includes gastroenteritis, eye, ear and skin infections, malaria, yellow fever, schistosomiases, dengue fever, diarrhea, dysentery, pyogenic infections, dyspepsia and urinary tract infections. (Bharti *et al*., 2003). The high prevalence of diarrhea among children and toddlers can be traced to the usage of polluted water and unhygienic practices (Tortora *et al*., 2002). Drinking water is also the most important source of gastro enteric diseases worldwide, mainly due to the fecal contamination of raw water, failure in the water treatment process or recontamination of drinking water at source and point of use (WHO, 2010). The major sources of faecal pollution of water and water borne *E.coli* infections are wild and farm animals feeding in water catchments (Chalmers *et al*., 2000). Toxic substances from manufacturing and agricultural practices leached from the land move into water in abundant quantity and they could be recalcitrant. Also, rural water frequently have excess nitrite, derived from microbial action on agricultural fertilizers (Izah and Ineyougha, 2017). The quality of water can be deteriorated in the course of collection, transport, and home storage. Thus, access to a safe source alone does not guarantee the quality of water that is consumed. To achieve a safe water supply to various societies, an understanding of water that is microbiologically and chemically certified is therefore vital. (WHO, 2010). Therefore, the aim of the research is to comparatively analyze the bacterial load and physicochemical characteristics of water samples from various sources used by the students of Federal University, Dutsinma Katsina State to determine their portability for ingestion and other domestic uses.

## MATERIALS AND METHODS

### The Study Area

Dutsin-Ma LGA lies on latitude 12°26’N and longitude 07°29’E. It is bounded by Kurfi and Charanchi LGAs to the north, Kankia LGA to the east, Safana and Dan-Musa LGAs to the west, and Matazu LGA to the southeast. Dutsin-Ma LGA has a land size of about 552.323 km2 with a population of 169 829 as at 2006 national census (Federal Republic of Nigeria, 2012). The people are predominantly farmers, pastoral farmers and traders. The climate of Katsina State is the tropical wet and dry type (tropical continental climate) Rainfall is between May and September with a peak in August. The average annual rainfall is about 700 mm.

### Sample collection

Samples of tap, well, dam, rain, sachet and boreholes were collected at different locations where students reside. For samples collected from wells and dam, the sample bottles were filled from below the surface of the water to avoid floating particles. While samples collected from taps and boreholes were taken after allowing the tap to run for about five minutes. The plastic bottles were caped, placed in an icepack and transported to the Department of Microbiology, Federal University Dutsinma, for further analysis. There were 6 sources of water, namely; tap water, dam water, well water, borehole water, sachet water and rain water from which 10 samples were obtained each, making a total of 60 samples.

### Bacterial count from water sample

The samples were serially diluted, 9ml of distilled water was pipette into 3 sterile test tubes. 1ml of the water sample was pipette into the first test tube and was shaken vigorously to have a homogeneous mixture (stock). 1ml of the mixture was pipette into the second test tube containing distilled water and from the second test tube 1ml was pipette into the last test tube. The steps were repeated to the remaining samples until a dilution of 10^3^ is obtained. 1ml of the dilution was poured plated into sterile petri dishes containing nutrient agar to determine the bacterial load count; the plates were incubated at 37°c for 24 hours (Chessbrough, 2000)

### Physiochemical analysis

Physical analysis was first carried out. A little quantity of the water sample was poured into the beaker and the temperature, pH and electrical conductivity were taken at the site of collection.

### Enumeration of Total bacteria count

Serial dilution of water sample was done using 9ml of distilled water and 1ml of water sample for each of the water sample and was weighed in the test tube. Poured plate method was used, using nutrient agar and was incubated at 35°C for 24 hours, then colonies were counted. Three plates each were used for a particular water sample. The average number of colonies from the three plates was taken and recorded. Water of good quality has a low total bacterial count fewer than 100cfu per millimeters (Chessbrough, 2000).

### Cultural characteristics and identification of isolates

Each colony and morphology e.g. size, margin, elevation, color, transparency was determined. Identification of isolates were done based on microscopic appearance (Gram stain) and biochemical test.

## BIOCHEMICAL TEST

Biochemical test was followed after the segregation of unadulterated culture from various agar media, the way of life were then protected and were later exposed to different biochemical tests for the confirmation and identification of the disengages. The biochemical tests did were: Catalase test, Coagulase test, citrate test, indole test, urease test, Triple sugar iron test, motility test and oxidase test.

## RESULTS

Table 1 shows the physicochemical properties of water quality from different sources within several locations in Dutsin-ma. Seven physicochemical parameters were analyzed. These are pH, temperature, turbidity, electrical conductivity, dissolved oxygen, hardness and total dissolved solids respectively.

**Table 1.**
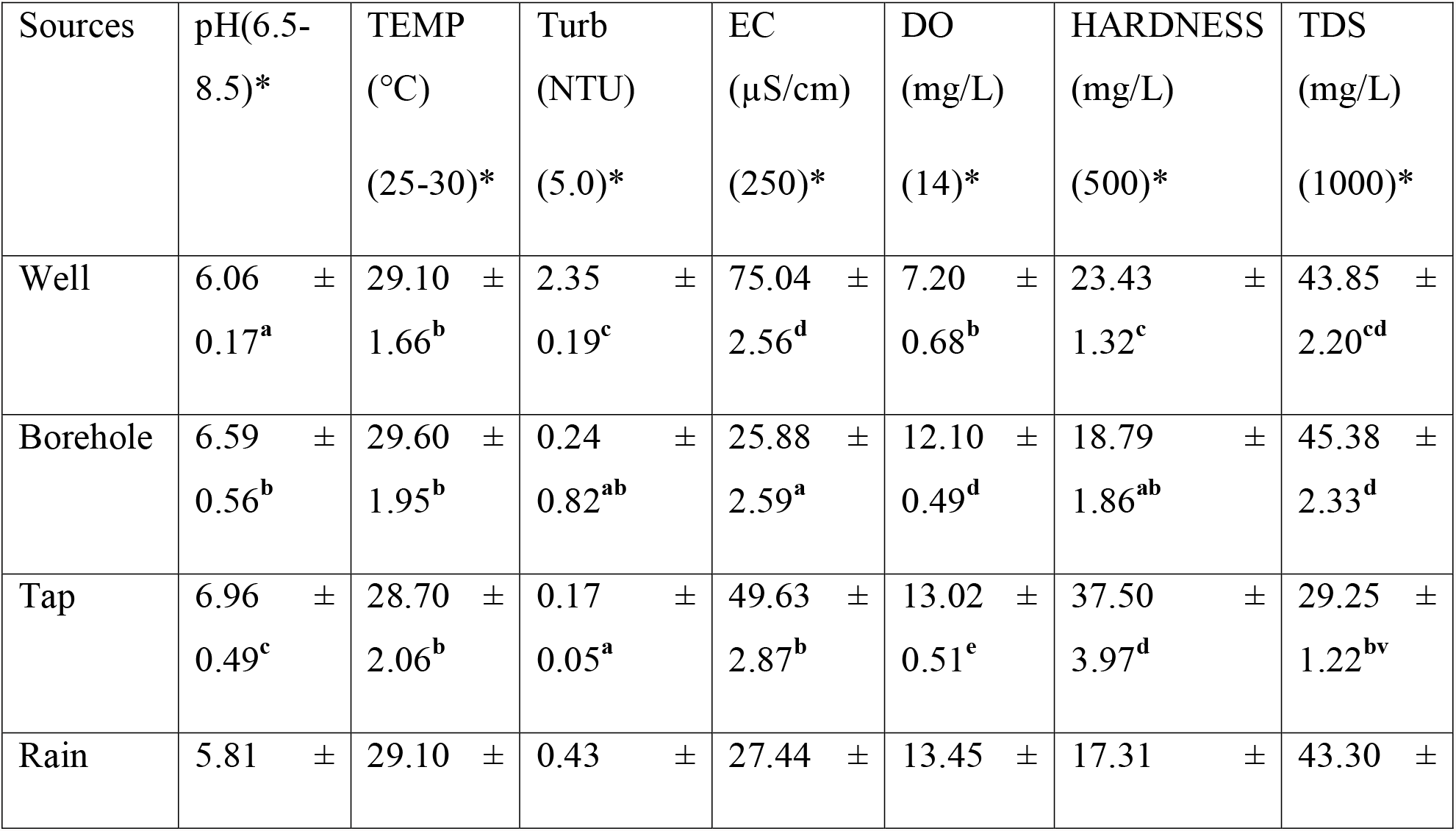
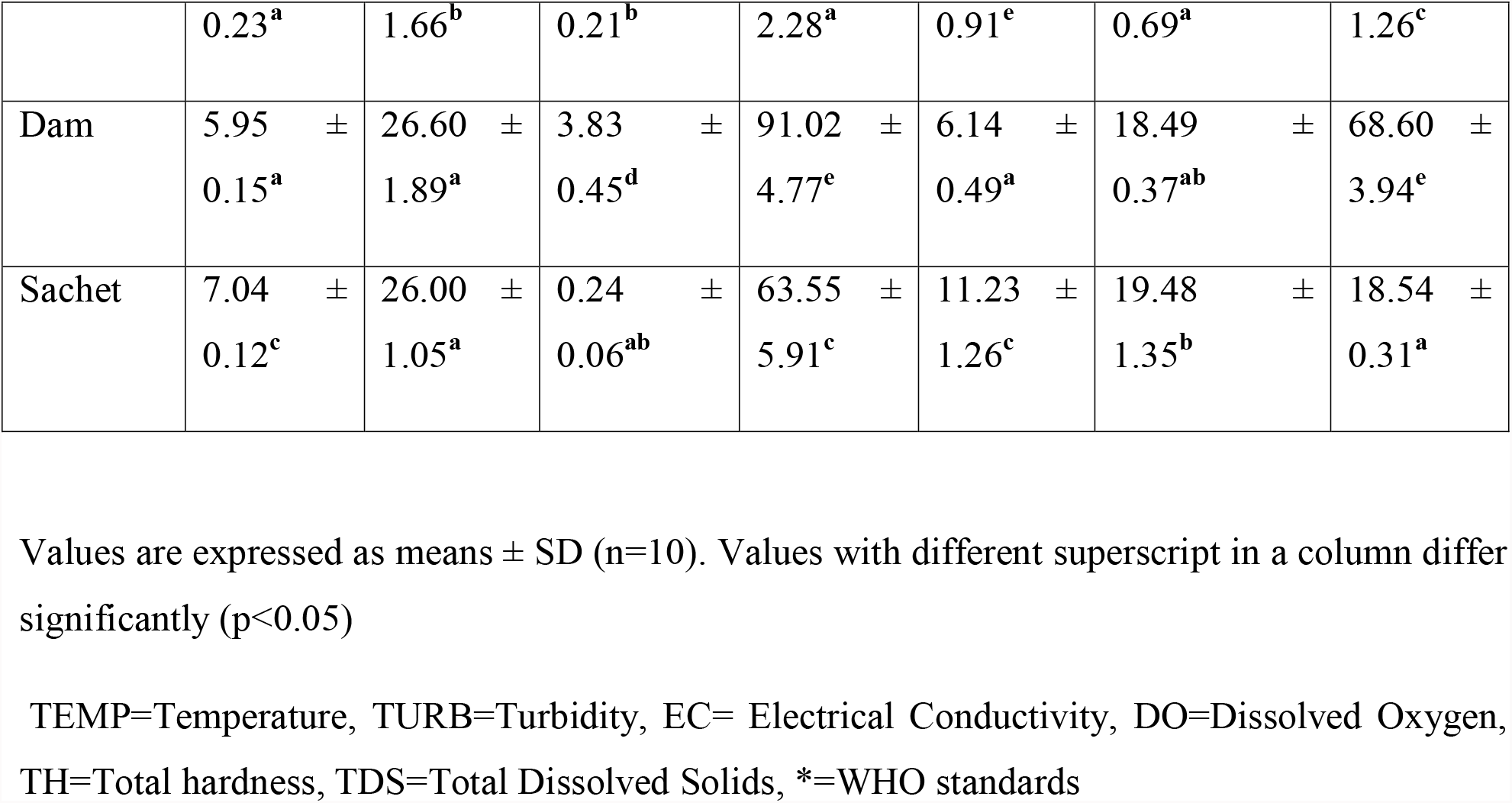
Physiochemical properties of water quality from different sources.

### Total plate count for the organisms in borehole water samples DF =0

Table 2 shows the total plate count for the organisms in borehole water samples which represents the mean number of bacteria colonies found in each triplicate.

**Table 2.** Total plate count for the organisms in borehole water samples DF =0.

| SAMPLES | AVERAGE | VOLUME | STANDARD PLATE |
| --- | --- | --- | --- |
|  | COLONIES | PLATED(ml) | COUNT( CFU/ml) |
| B1 | 45 | 1 | $4.5 \times 10^1$ |
| B2 | 43 | 1 | $4.3 \times 10^1$ |
| B3 | 48 | 1 | $4.8 \times 10^1$ |
| B4 | 41 | 1 | $4.1 \times 10^1$ |
| B5 | 45 | 1 | $4.5 \times 10^1$ |
| B6 | 44 | 1 | $4.4 \times 10^1$ |
| B7 | 49 | 1 | $4.9 \times 10^1$ |
| B8 | 53 | 1 | $5.3 \times 10^1$ |
| B9 | 46 | 1 | $4.6 \times 10^1$ |
| B10 | 42 | 1 | $4.2 \times 10^1$ |

**KEY: B1=Borehole water sample 1, B2= Borehole water sample 2, B3= Borehole water sample 3, B4= Borehole water sample 4, B5= Borehole water sample 5, B6= Borehole water sample 6, B7= Borehole water sample 7, B8= Borehole water sample 8, B9= Borehole water sample 9, B10= Borehole water sample 10.**

### Total plate count for the organisms in well water samples DF=10^3^

Table 3 shows the total plate count for the organisms in well water samples which represents the mean number of bacteria colonies found in each triplicate.

**Table 3.**
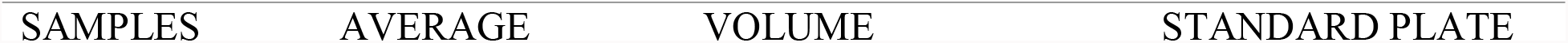
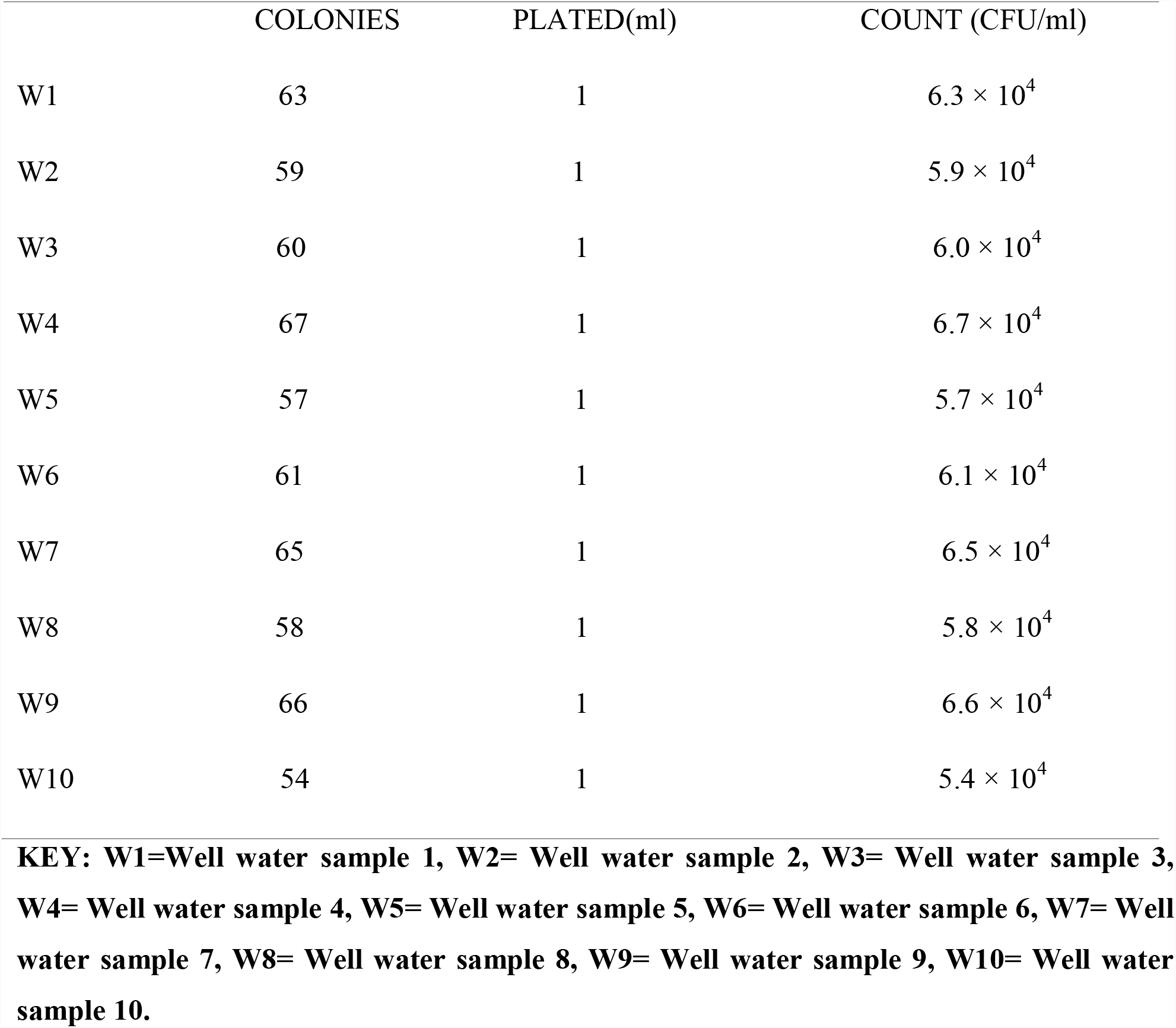
Total plate count for the organisms in well water samples DF=10^3^.

Table 4 shows the total plate count for the organisms in dam water samples which represents the mean number of bacteria colonies found in each triplicate.

**Table 4.** Total plate count for the organisms in dam water samples DF=10^3^.

| SAMPLES | AVERAGE | VOLUME | STANDARD PLATE |
| --- | --- | --- | --- |
|  | COLONIES | PLATED(ml) | COUNT (CFU/ml) |
| D1 | 86 | 1 | $8.6 \times 10^4$ |
| D2 | 79 | 1 | $7.9 \times 10^4$ |
| D3 | 87 | 1 | $8.7 \times 10^4$ |
| D4 | 89 | 1 | $8.9 \times 10^4$ |
| D5 | 83 | 1 | $8.3 \times 10^4$ |
| D6 | 85 | 1 | $8.5 \times 10^4$ |
| D7 | 90 | 1 | $9.0 \times 10^4$ |
| D8 | 79 | 1 | $7.9 \times 10^4$ |
| D9 | 80 | 1 | $8.0 \times 10^4$ |
| D10 | 91 | 1 | $9.1 \times 10^4$ |

**KEY: D1=Dam water sample 1, D2= Dam water sample 2, D3= Dam water sample 3, D4= Dam water sample 4, D5= Dam water sample 5, D6= Dam water sample 6, D7= Dam water sample 7, D8= Dam water sample 8, D9= Dam water sample 9, D10= Dam water sample 10.**

Table 5 shows the total plate count for the organisms in borehole water samples which represents the mean number of bacteria colonies found in each triplicate.

**Table 5.**
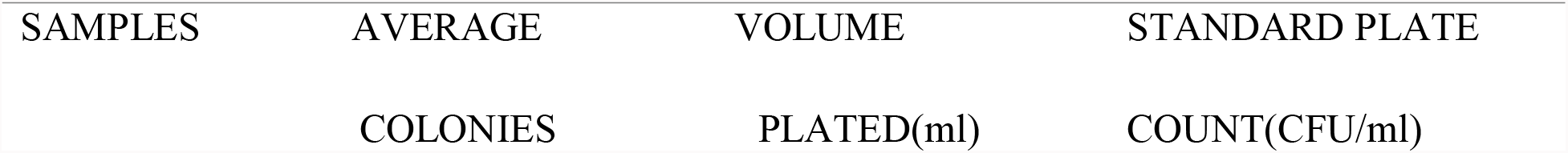
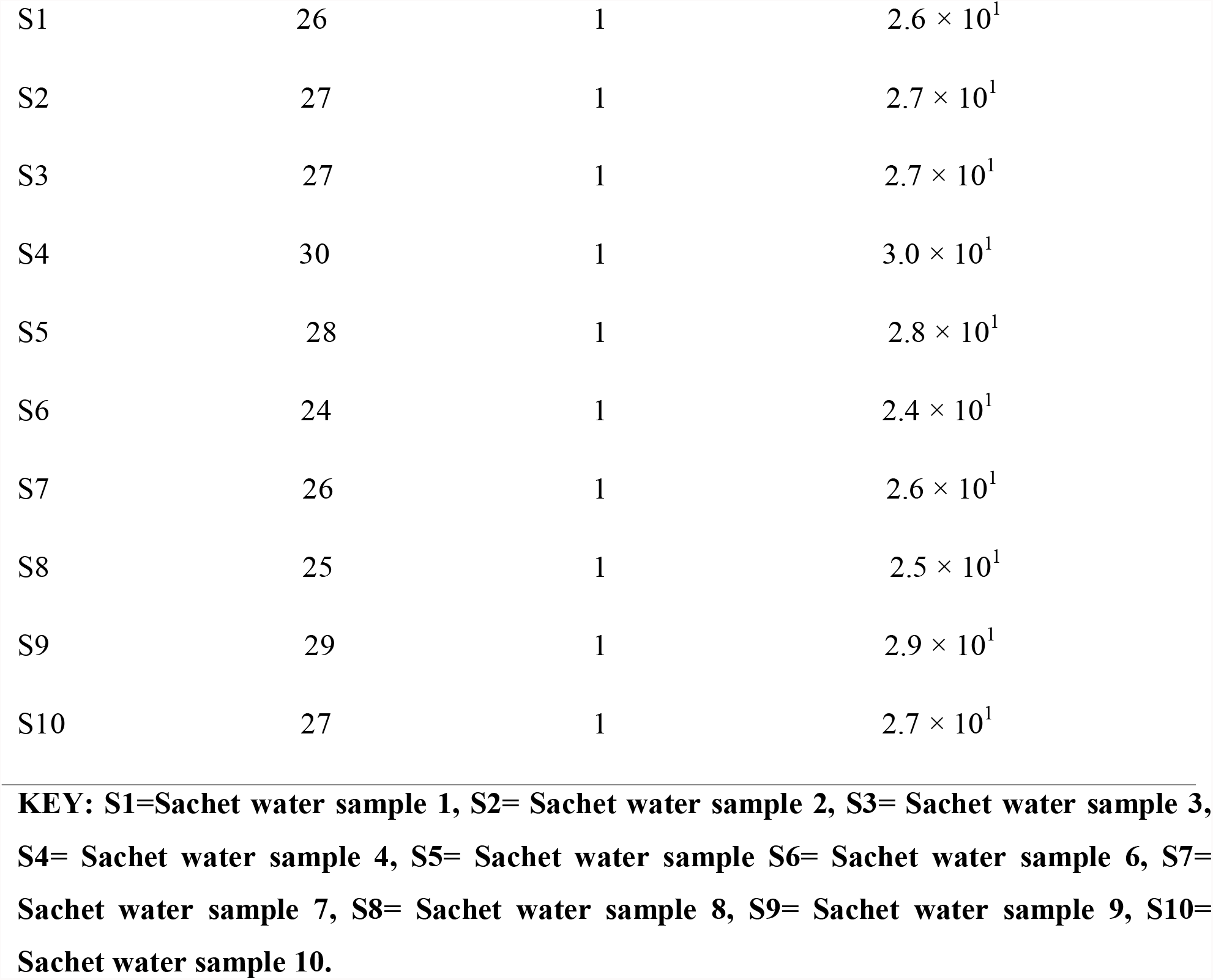
Total plate count for the organisms in sachet water samples DF=0.

| SAMPLES | AVERAGE | VOLUME | STANDARD PLATE |
| --- | --- | --- | --- |
|  | COLONIES | PLATED(ml) | COUNT(CFU/ml) |
| S1 | 26 | 1 | $2.6 \times 10^1$ |
| S2 | 27 | 1 | $2.7 \times 10^1$ |
| S3 | 27 | 1 | $2.7 \times 10^1$ |
| S4 | 30 | 1 | $3.0 \times 10^1$ |
| S5 | 28 | 1 | $2.8 \times 10^1$ |
| S6 | 24 | 1 | $2.4 \times 10^1$ |
| S7 | 26 | 1 | $2.6 \times 10^1$ |
| S8 | 25 | 1 | $2.5 \times 10^1$ |
| S9 | 29 | 1 | $2.9 \times 10^1$ |
| S10 | 27 | 1 | $2.7 \times 10^1$ |

**KEY: S1=Sachet water sample 1, S2= Sachet water sample 2, S3= Sachet water sample 3, S4= Sachet water sample 4, S5= Sachet water sample S6= Sachet water sample 6, S7= Sachet water sample 7, S8= Sachet water sample 8, S9= Sachet water sample 9, S10= Sachet water sample 10.**

Table 6 shows the total plate count for the organisms in rain water samples which represents the mean number of bacteria colonies found in each triplicate.

**Table 6.**
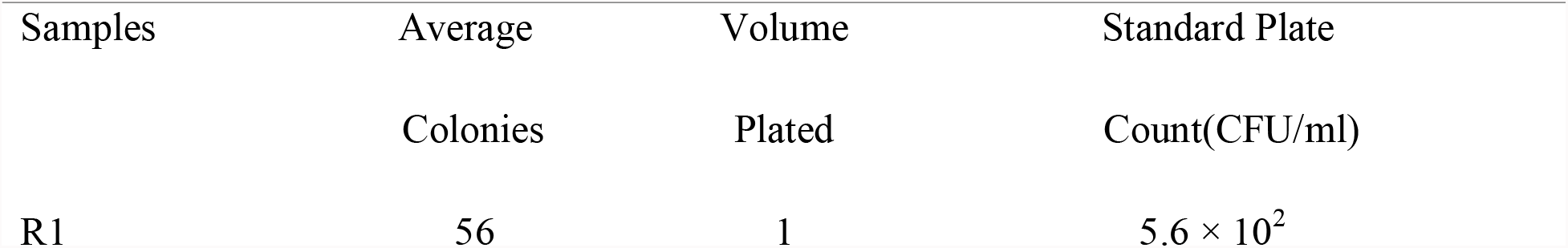
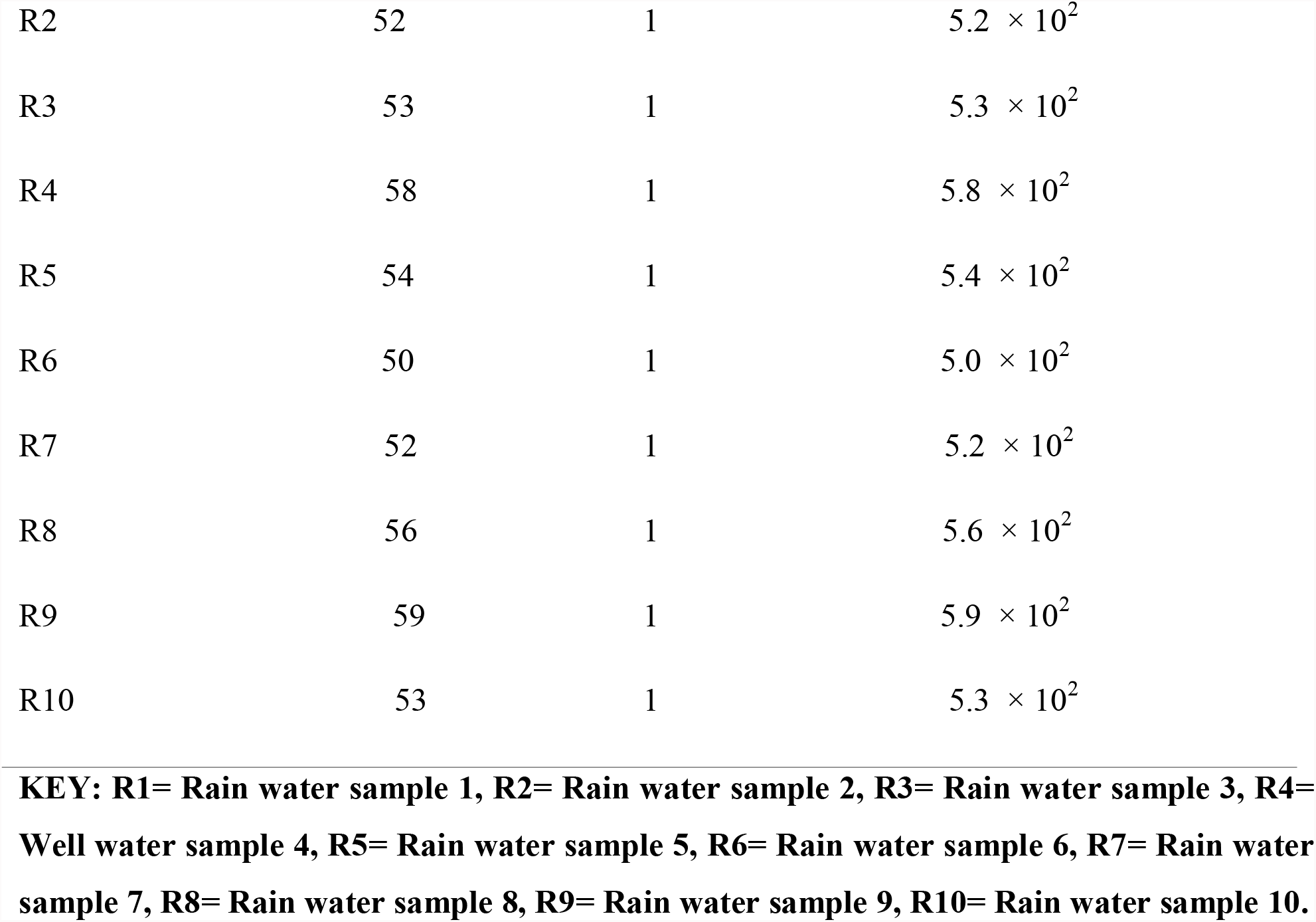
Total plate count for the organisms in rain water samples DF=10^1^.

| Samples | Average | Volume | Standard Plate |
| --- | --- | --- | --- |
|  | Colonies | Plated | Count(CFU/ml) |
| R1 | 56 | 1 | $5.6 \times 10^2$ |
| R2 | 52 | 1 | $5.2 \times 10^2$ |
| R3 | 53 | 1 | $5.3 \times 10^2$ |
| R4 | 58 | 1 | $5.8 \times 10^2$ |
| R5 | 54 | 1 | $5.4 \times 10^2$ |
| R6 | 50 | 1 | $5.0 \times 10^2$ |
| R7 | 52 | 1 | $5.2 \times 10^2$ |
| R8 | 56 | 1 | $5.6 \times 10^2$ |
| R9 | 59 | 1 | $5.9 \times 10^2$ |
| R10 | 53 | 1 | $5.3 \times 10^2$ |

**KEY: R1= Rain water sample 1, R2= Rain water sample 2, R3= Rain water sample 3, R4= Well water sample 4, R5= Rain water sample 5, R6= Rain water sample 6, R7= Rain water sample 7, R8= Rain water sample 8, R9= Rain water sample 9, R10= Rain water sample 10.**

Table 7 shows the total plate count for the organisms in borehole water samples which represents the mean number of bacteria colonies found in each triplicate.

**Table 7.** Total plate count for the organisms in tap water samples DF=10^1^.

| SAMPLES | AVERAGE | VOLUME | STANDARD PLATE |
| --- | --- | --- | --- |
|  | COLONIES | PLATED | COUNT (CFU/ml) |
| T1 | 55 | 1 | $5.5 \times 10^2$ |
| T2 | 51 | 1 | $5.1 \times 10^2$ |
| T3 | 41 | 1 | $4.1 \times 10^2$ |
| T4 | 48 | 1 | $4.8 \times 10^2$ |
| T5 | 45 | 1 | $4.5 \times 10^2$ |
| T6 | 49 | 1 | $4.9 \times 10^2$ |
| T7 | 47 | 1 | $4.7 \times 10^2$ |
| T8 | 42 | 1 | $4.2 \times 10^2$ |
| T9 | 43 | 1 | $4.3 \times 10^2$ |
| T10 | 46 | 1 | $4.6 \times 10^2$ |

**KEY:** T1= Tap water sample 1, T2= Tap water sample 2, T3= Tap water sample 3, T4= Tap water sample 4, T5= Tap water sample 5, T6= Tap water sample 6, T7= Tap water sample 7, T8= Tap water sample 8, T9= Tap water sample 9, T10= Tap water sample 10.

Table 8 shows the frequency of occurrence and percentage of bacteria found in each source of water. Bacteria count of each water sample was carried out and presence of *Escherichia coli, P.aeruginosa, S.aureus, S*.typhi, *K.pneumoniae, B.subtilis, Proteus sp, Shigella sp*, and *E.aerogenes* were identified using pour plate method on suitable media as described by Chessbrough, (2000).

**Table 8.**
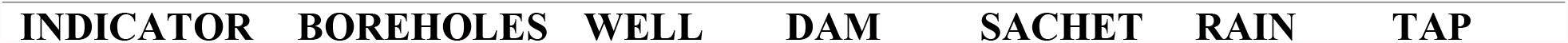
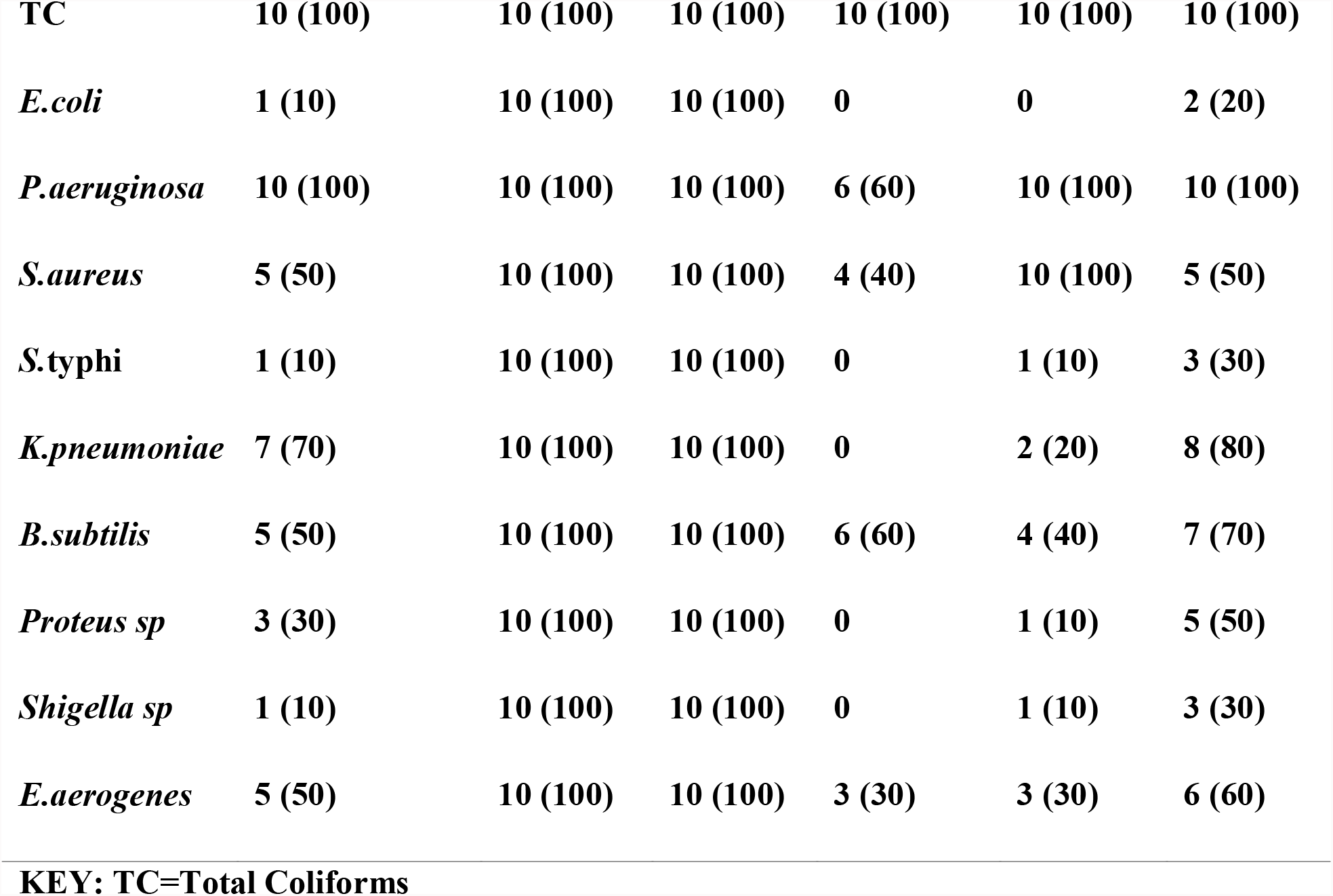
Prevalence of lactose fermenting bacteria (coliforms) bacteria, indicator organisms and detection of *Escherichia coli* on eosin methylene blue agar.

| INDICATOR | BOREHOLES | WELL | DAM | SACHET | RAIN | TAP |
| --- | --- | --- | --- | --- | --- | --- |
| **TC** | **10 (100)** | **10 (100)** | **10 (100)** | **10 (100)** | **10 (100)** | **10 (100)** |
| *E.coli* | **1 (10)** | **10 (100)** | **10 (100)** | **0** | **0** | **2 (20)** |
| *P.aeruginosa* | **10 (100)** | **10 (100)** | **10 (100)** | **6 (60)** | **10 (100)** | **10 (100)** |
| *S.aureus* | **5 (50)** | **10 (100)** | **10 (100)** | **4 (40)** | **10 (100)** | **5 (50)** |
| *S.typhi* | **1 (10)** | **10 (100)** | **10 (100)** | **0** | **1 (10)** | **3 (30)** |
| *K.pneumoniae* | **7 (70)** | **10 (100)** | **10 (100)** | **0** | **2 (20)** | **8 (80)** |
| *B.subtilis* | **5 (50)** | **10 (100)** | **10 (100)** | **6 (60)** | **4 (40)** | **7 (70)** |
| *Proteus sp* | **3 (30)** | **10 (100)** | **10 (100)** | **0** | **1 (10)** | **5 (50)** |
| *Shigella sp* | **1 (10)** | **10 (100)** | **10 (100)** | **0** | **1 (10)** | **3 (30)** |
| *E.aerogenes* | **5 (50)** | **10 (100)** | **10 (100)** | **3 (30)** | **3 (30)** | **6 (60)** |

**KEY: TC=Total Coliforms**

Table 15 shows the biochemical characteristics of the bacteria isolated from the water samples. Among the bacteria isolated, *E.coli* was found to be the most frequent and highest occurring organism.

**TABLE 15:** BIOCHEMICAL CHARACTERISTICS OF ISOLATE.

| ISOL | CA | CIT | COA | MO | IN | OX | URE | TSI | ORGANISM |
| --- | --- | --- | --- | --- | --- | --- | --- | --- | --- |
| ATE |  |  |  |  |  |  |  |  |  |
| A | + | + | – | – | – | + | – | ALKALINE/ALKALINE | *P.aeruginosa* |
| B | + | + | + | - | - | - | + | - | *S.aureus* |
| C | + | - | - | + | - | - | - | ALKALINE/ACID | *S. TYPHI* |
|  |  |  |  |  |  |  |  | H<sub>2</sub>S PRODUCED |  |
| D | + | + | - | - | - | - | + | ACID/ACID | *K. pneumoniae* |
|  |  |  |  |  |  |  |  | GAS PRODUCED |  |
| E | + | + | - | + | - | - | - | - | *B.subtilis* |
| F | + | + | - | - | + | - | - | - | *Proteus sp* |
| G | + | - | - | - | + | - | - | ALKALINE/ACID | *Shigella sp* |
| H | + | + | - | - | + | - | - | ACID/ACID | *E.aerogenes* |
|  |  |  |  |  |  |  |  | GAS PRODUCED |  |
| I | + | - | - | + | + | - | - | ACID/ACID | *E.coli* |
|  |  |  |  |  |  |  |  | GAS PRODUCED |  |

**KEY: CA=Catalase Test, CIT=Citrate Test, COA=Coagulase Test, MO=Motility Test, IN=Indole Test, OX=Oxidase Test, URE=Urease Test, TSI=Triple Sugar Iron Test.**

## DISCUSSION

The research reveals the microbial quality of water sources used by students of Federal University Dutsin-ma. The research found that the microbial quality of most water samples exceed World Health Organization allowable limit of 1.0 × 10^2^ cfu/ml for potable water and Standard Organization of Nigeria maximum permissible level of 10cfu/ml (total coliform) and 0 cfu/100ml (Thermo tolerant Coliform or *E. coli*). However, surface water has high microbial load than the ground water. The microbial populations are typically highest in surface water and ground water (well water □ground water), followed by rain water and least in sachet. All the sachet water used for drinking purposes contains coliforms. These coliforms may not be harmful to people but it implies that the treatment process used by water processing industries is not working properly. The microbial density i.e total heterotrophic bacteria count; total coliform, fecal coliform exceeds the recommended limits. Typically, microbial load of the different portable water sources in the order are surface water (Dam) > well □ tap□ borehole □ rain □ sachet water. However, microbial density were least in sachet water. Several dominant microbial diversity found in the potable water sources including *Staphylococcus aureus, Escherichia* coli, *Proteus, Pseudomonas, Enterobacter, Salmonella, Klebsiella, Bacillus* species e.t.c are known to cause diseases conditions. The pH range (5.8 -6.3) for well water, (5.5 – 6.2) for rain water and (5.7 -6.2) for Dam water could be considered as being an unacceptable range for natural water. According to Okonko *et al*., (2008), the pH of most natural waters range from 6.5 – 8.5. The low PH values obtained in Dam water might be due to the high levels of free CO_2_ in the water samples, which might affect the bacterial count. This was also reported by Edema *et al*. (2011).The PH of water is extremely important. The fluctuations in optimum PH ranges may lead to an increase or decrease in in the toxicity of poisons in water bodies (okonko *et al*., 2008). Long term exposure to pH beyond the allowable limit affects the mucous membrane of cells (Nishtha *et al*. 2012). The water from the borehole groundwater sources and tap water that were found to be acidic can also easily corrode the water piping due to the acidic nature of the water. Damaged metal pipes due to acidic pH values can also lead to problems, causing water to have a metallic or sour taste (Sabrina *et al*. 2013). The highest turbidity was recorded in the Dam water source and the lowest turbidity was recorded in the sachet water. The turbidity of water sources in order are surface water (Dam water > well water □ tap water □borehole □ rain water □sachet water). All the samples analyzed were found within the limits prescribed by WHO standards (1– 5 NTU). Turbidity indicates that there may be the presence of inorganic particulate matter and non-soluble metal oxides. The intake of high turbid water may cause a health risk, as excessive turbidity can protect pathogenic microorganisms from effects of disinfectants (Singh *et al*. 2013; Tiwari and Singh 2014). Total dissolved solids indicate the salinity behavior of groundwater. In the study area, TDS values varied from 18.11 mg/L to 75.31 mg/L. All the samples analyzed were found within the standard permissible limit. The TDS of water sources in order are (Dam water > well water □ tap water □borehole □ rain water □sachet water). The presence of TDS above limit in water sources would cause undesirable taste and gastrointestinal irritation (Selvakumar *et al*. 2014). EC means the conducting capacity of water. EC is a measure of total dissolved solids (TDS) i.e., it depends upon the ionic strength of the solution. Increase in the concentration of dissolved solids increases the ionic strength of the solution. In the study area, EC was in the range of 53.1µS/cm to 98.2µS/cm. The EC of water sources in order are surface water (Dam)> well □tap □borehole□rain □sachet water. All the samples analyzed were found within the limits prescribed by WHO standards (500µS/cm). TDS and EC increases the acidic nature of water, unusual taste, odor and feel problems usually due to total dissolved solids and higher EC indicate the presence of dissolved minerals (WHO 2010). The hardness of water samples in the study area varied from 17.2 to 41.9 mg/L. All the samples analyzed were found within the limits prescribed by WHO standards (500mg/L). High hardness may cause deposit on water supply distribution systems, hardness above 100 mg/l may result in the need for more soap during bathing and laundering; forms scum; causes yellowing of fabrics; toughens vegetables cooked in the water; excessive hardness may also lead to scale deposits in pipes, heaters, and boilers. Long term consumption of extremely hard water might lead to an increased incidence of urolithiasis, anencephaly, prenatal mortality, some types of cancer and cardiovascular disorders (Agrawal and Jagetia 1997). For Dissolved Oxygen, All the samples analyzed were found within the limits prescribed by WHO standards (14 mg/L).The microbiological analysis of Dam and well water samples shows the highest microbial load which was higher than the recommended value. The presence of *Enterobacter sp* in sachet water may be due to poor storage while the presence of bacillus may be from processing and inadequate chemical usage. The presence of *S.aureus* is because it is highly commensal and ubiquitous.

## CONCLUSION

From the results, it shows that sachet water in Dutsinma is safe for consumption. The coliforms in tap water slightly exceeds the standards by WHO and two samples from tap water had fecal contaminations which makes it not portable for human consumption. Also, the study revealed that the distribution pattern of the studied physico–chemical parameters (except pH) of borehole water, sachet water, Dam, Rain, well, tap water were within the permissible limit set by World Health Organization. Only sachet water is fit for consumption. Rain water has less bacterial load but has an acidic pH, therefore it is unfit for consumption. Tap and borehole water should be treated before consumption. Dam water and well water can be used for other domestic purposes.

## Data Availability

All data produced in the present study are available upon reasonable request to the authors

## RECOMMENDATIONS

The inhabitants in Dutsinma should be educated on the potential implications and economic importance of drinking contaminated water.

The government should provide portable drinking water that meets the national and international drinking water standards by thorough monitoring of water purification.

Sachet water should be appropriately stored and the air quality of the surroundings should be bacteriologically tested. The environment should also be monitored frequently.

Government should dedicate a day or two only for Environmental sanitation periodically to lessen air blown pollutants into the Dam and well water. Water treatment systems should be designed to eradicate harmful organism that pose a health risk to man.

There should be a standard set aside for borehole water construction both for private and general (municipal) use/purposes.

Construction of soak away near a water source should be banned.

From the result obtained, water quality monitoring should be an incessant process that should be encouraged.

Further study should be carried out on carbon-based contaminants like pesticides, aldehydes, phenols and other toxic element such as mercury and arsenic.

NAFDAC should implement and ensure stern compliance to the standards as regard the production and sales of packaged water.

Well water should be boiled before consumption to ensure good health safety.

